# Switched and unswitched memory B cells detected during SARS-CoV-2 convalescence correlate with limited symptom duration

**DOI:** 10.1101/2020.09.04.20187724

**Authors:** Krista L. Newell, Deanna C. Clemmer, Justin B. Cox, Yetunde I. Kayode, Victoria Zoccoli-Rodriguez, Harry E. Taylor, Timothy P. Endy, Joel R. Wilmore, Gary M. Winslow

**Author notes:** Address correspondence to Dr. Gary M. Winslow, Upstate Medical University, 766 Irving Avenue, Syracuse, NY 13210. 1-315-464-7658.

## Abstract

Severe acute respiratory syndrome coronavirus-2 (SARS-CoV-2), the causative agent of the pandemic human respiratory illness COVID-19, is a global health emergency. While severe acute disease has been linked to an expansion of antibody-secreting plasmablasts, we sought to identify B cell responses that correlated with positive clinical outcomes in convalescent patients. We characterized the peripheral blood B cell immunophenotype and plasma antibody responses in 40 recovered non-hospitalized COVID-19 subjects that were enrolled as donors in a convalescent plasma treatment study. We observed a significant negative correlation between the frequency of peripheral blood memory B cells and the duration of symptoms for convalescent subjects. Memory B cell subsets in convalescent subjects were composed of classical CD24^+^ class-switched memory B cells, but also activated CD24-negative and natural unswitched CD27^+^ IgD^+^ IgM^+^ subsets. Memory B cell frequency was significantly correlated with both IgG1 and IgM responses to the SARS-CoV-2 spike protein receptor binding domain (RBD).

IgM^+^ memory, but not switched memory, directly correlated with virus-specific antibody responses, and remained stable over time. Our findings suggest that the frequency of memory B cells is a critical indicator of disease resolution, and that IgM^+^ memory B cells play an important role in SARS-CoV-2 immunity.

## Introduction

There have now been over 24 million reported cases of SARS-CoV-2, including at least 830,000 deaths worldwide (1). As the work to develop effective vaccines and therapies to control the pandemic progresses, it is important to develop reliable approaches for assessing durable immunological memory. Identification of a correlate of immunity to SARS-CoV-2 has been challenging, as clinical presentation and serological profiles vary between patients. Rare SARS-CoV-2-specific antibodies with potent neutralizing capacity have been isolated from recovered COVID-19 patients (2). Additionally, acute COVID-19 patients have been observed to have perturbations of immune profiles, and have been grouped into three or more immunotype clusters (3). On the basis of these findings, we focused on correlates of clinical outcomes in convalescent plasma donors that could be inferred through cell-based assays. Recent studies have correlated B cell responses in some individuals with immunity and protection (4).

B cells participate in the antiviral immune response by first rapidly releasing germline or near-germline antibodies from plasmablasts, via an extrafollicular pathway. Upon T cell-mediated CD40 ligation and appropriate cytokine stimulation, B cells undergo class switching and/or enter germinal centers within secondary lymphoid organs to undergo affinity maturation. This maturation process produces both long-lived plasma cells and memory B cells capable of responding to secondary challenge with homotypic or heterotypic antigenic challenge.

While many studies of the B cell response to SARS-CoV-2 have focused on plasmablast expansion, the benefit of this expansion has not been clear (3, 5, 6). Indeed, clinical outcome correlations suggest that extrafollicular B cell activation and subsequent plasmablast generation are detrimental to host survival and COVID-19 symptom resolution. Memory B cells are also generated following SARS-CoV-2 infection (4). B memory cells are found as multiple subsets, including canonical CD27^+^ class-switched memory B cells, but also activated CD24-negative and “innate-like” natural CD27^+^ IgD^+^ IgM^+^ subsets. Increased memory B cell frequencies can reveal a successful response to acute viral infection, and can provide information regarding the quality of T cell help during the acute immune response. In this study, we monitored memory B cell subsets and their relationship to clinical parameters in convalescent COVID-19 subjects. We provide evidence of stable B cell memory populations in recovered subjects that correlate with attenuated symptom duration. We propose that a well-developed memory B cell response provides a reliable measure of immunity that may also be useful for evaluating SARS-CoV-2 vaccine efficacy.

## Results

### Longitudinal sampling of plasma donors with previous SARS-CoV-2 infection

To assess clinically-informative phenotypic characteristics of the B cell response in convalescent plasma donors, we evaluated peripheral blood PBMCs from this cohort and healthy volunteers by flow cytometry. Forty convalescent donors were recruited through the State University of New York Upstate Medical University Clinical Research Unit. They were sampled at an average of 69 days post-symptom onset, were a mean age of 51.6 years, and were composed of 35% males and 65% females. Donors were predominately white and non-Hispanic **(supplemental Figure 1)**. Convalescent donors were recruited following a positive SARS-CoV-2 PCR and at least 2 weeks following the last symptom, or for asymptomatic subjects, after a repeat SARS-CoV-2 PCR test that was negative. Seventeen subjects were also sampled at a 3-month follow-up visit. This latter subset included asymptomatic subjects, as well as those sampled in early, mid, and late convalescence. Characteristics of the study population are summarized in **Figure 1**.

**Figure 1.**
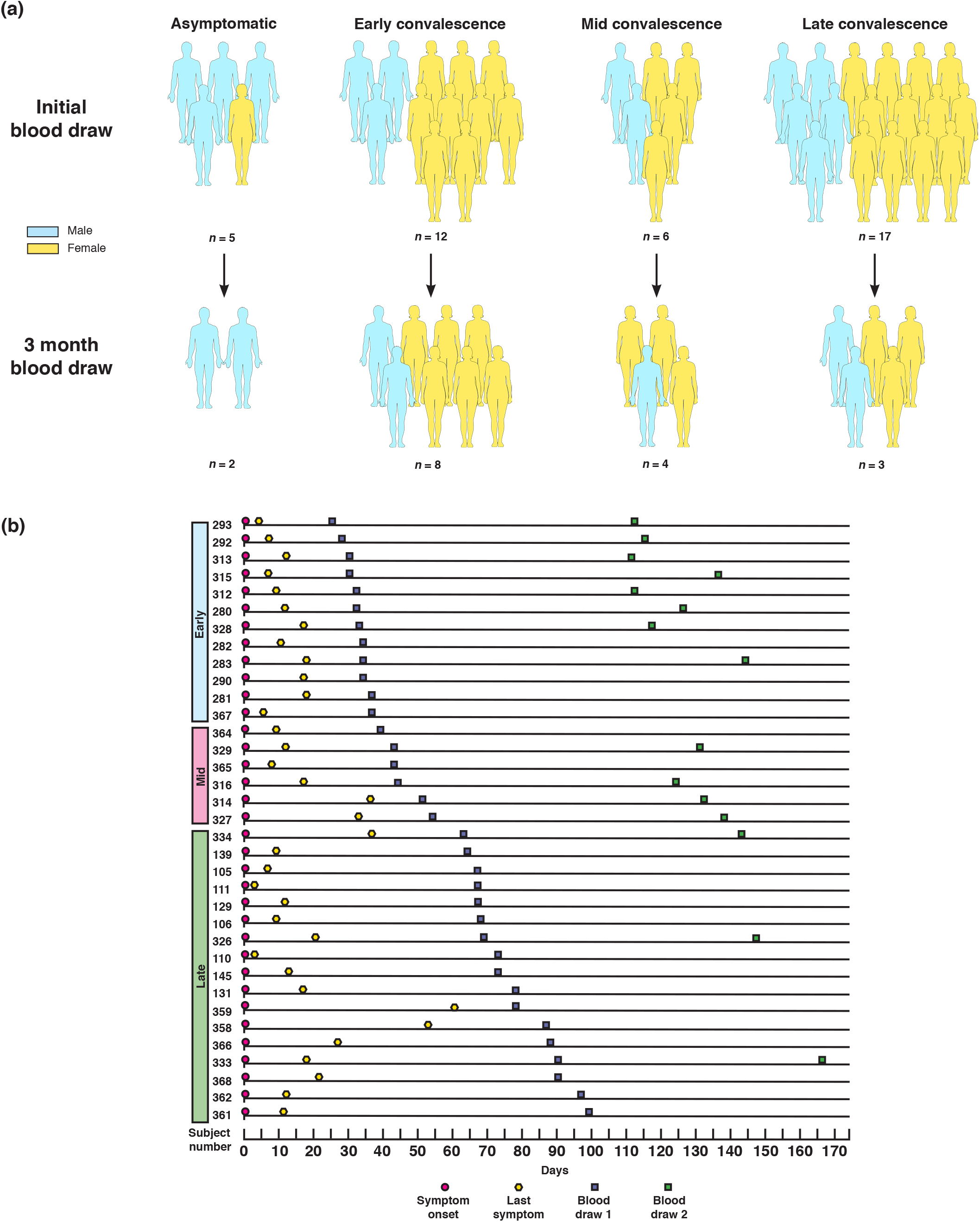
Study design and clinical data. **(a)** Graphical representation of convalescent subject groupings and gender. All grouping and subset designations were done retrospectively. **(b)** Symptom and sampling timeline for symptomatic subjects (n = 35), ordered by length of convalescence.

### B cell profiles varied widely during SARS-CoV-2 convalescence both between and within individuals

As has been observed in other studies (2, 5), the B cell profiles of convalescent plasma donors were diverse. Using a well-defined gating strategy described by Sanz et al. (7) **(supplemental Figure 2)**, we identified convalescent subjects with expanded B cell memory, a robust plasmablast population, and a B cell phenotype resembling that of many healthy control samples **(Figure 2a)**. We did not observe a difference in total CD19^+^ B cell frequency between convalescent and healthy subjects **(Figure 2b)**, but convalescent naïve, transitional, and activated B cell frequencies followed the inverse trend of memory frequencies over time **(Figure 2c-f)**. Memory B cell frequencies ranged widely, in both convalescent subjects and healthy controls **(Figure 2d)**. The plasmablast compartment was the only major B cell subset analyzed that remained significantly skewed in the convalescent phase compared to healthy controls **(Figure 2b-g)**. Moreover, elevated plasmablast frequencies were only observed in a subset of subjects.

**Figure 2.**
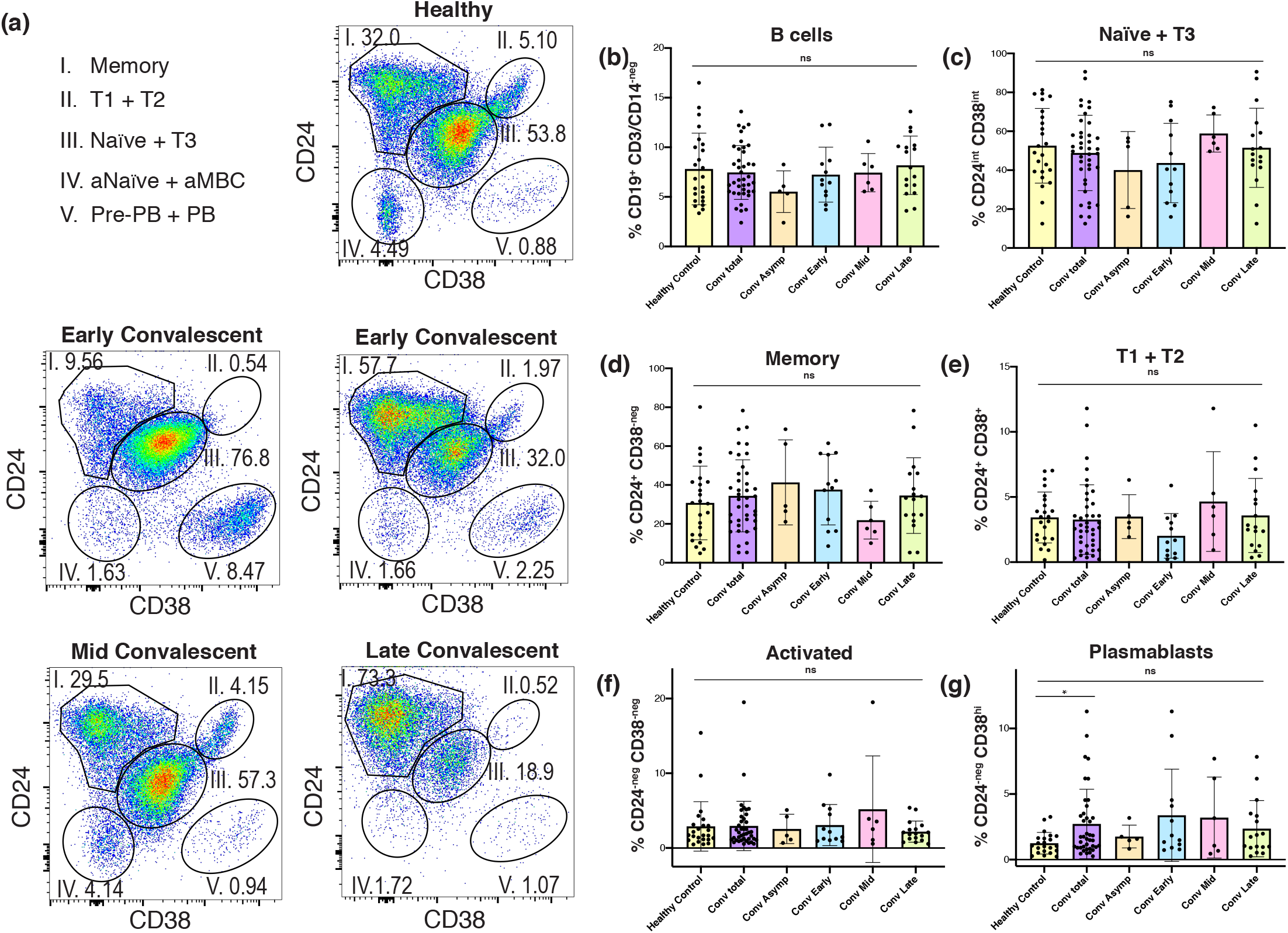
The B cell profiles of individual convalescent plasma donors were diverse. **(a)** Representative flow plots of live, singlet, CD19^+^ lymphocytes demonstrating 5 primary B cell subsets in one healthy and 4 convalescent subjects. **(b-d)** Histograms of healthy vs. convalescent donor frequencies of **(b)** total CD19^+^ B cells, and **(c)** naïve, **(d)** memory, **(e)** transitional 1 and 2, **(f)** activated naïve/memory, and **(g)** plasmablast subsets among viable CD19^+^ lymphocytes. Bars represent mean ^+^/-SD. Multiple comparison analysis between each donor subgroup was done with Kruskal-Wallis test with Dunn’s correction. Adjusted p value was used to determine family-wise significance at alpha = 0.05. Healthy control and total convalescent groups also compared by Mann-Whitney test with two-tailed p value, alpha = 0.05. Healthy control *n* = 24, conv. total *n* = 40, asymp. *n* = 5, conv. early *n* = 12, conv. mid *n* = 6, conv. late *n* = 17.

To further examine memory B cells in convalescent subjects, we visualized the flow cytometry datasets using an unbiased t-distributed stochastic neighbor embedding (t-SNE) algorithm **(Figure 3a)**. This approach allowed us to resolve populations within clusters that may not be discrete using gating strategies alone, and assess marker expression within these clusters. The majority of convalescent subjects had normal to elevated frequencies of both switched and unswitched CD38-negative CD24^+^ memory B cells **(Figure 3b-c)**. These data contrast with observations of memory B cell loss in COVID-19 patients with acute disease (8). The peripheral blood of convalescent subjects also contained a modest pool of activated CD38-negative CD24-negative B cells, which were the primary clusters in which CD11c and T-bet were expressed **(Figure 3a)**.

**Figure 3.**
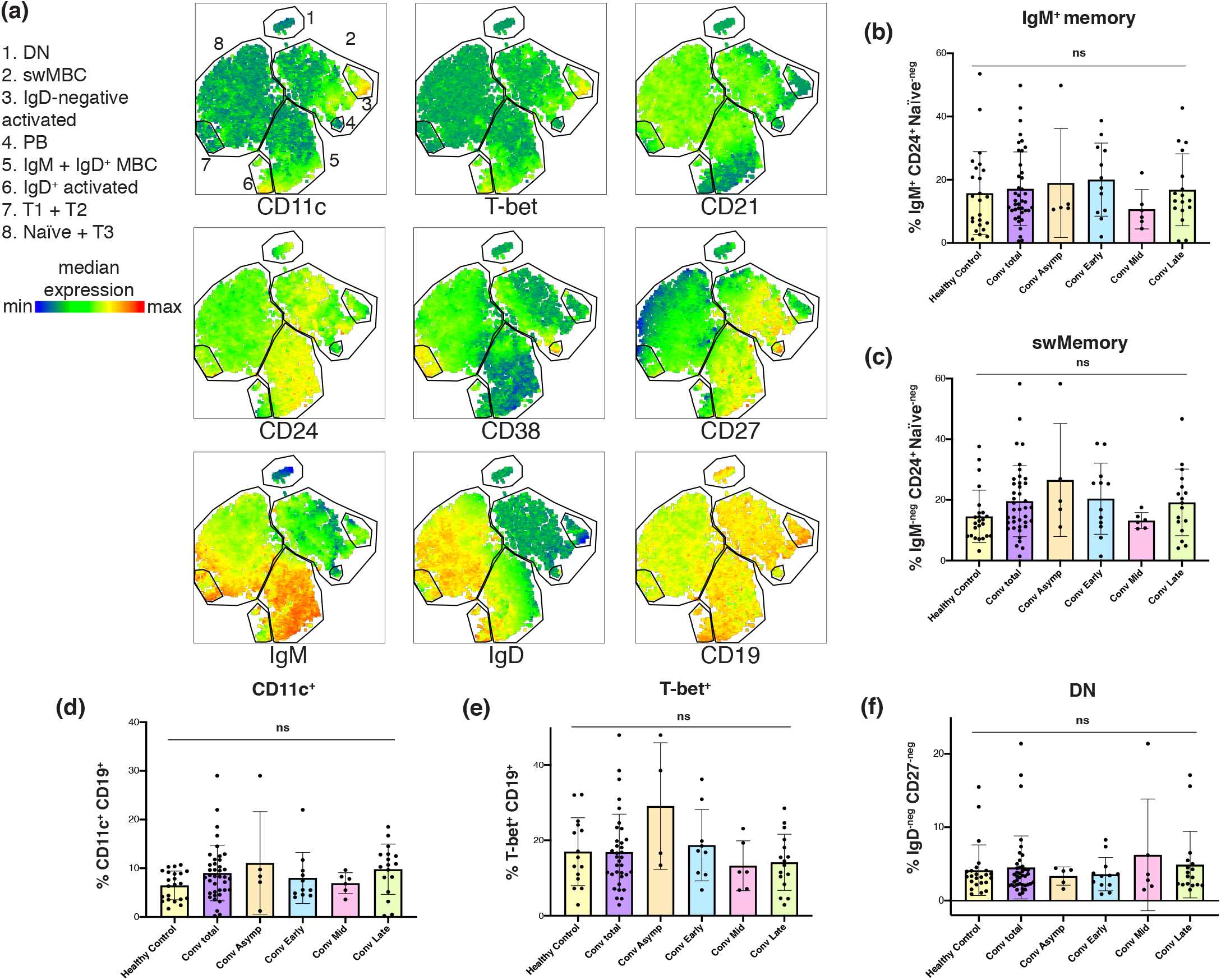
Convalescent plasma donor B cell compartments were heterogeneous and enriched for memory B cells. **(a)** tSNE representation sub-gated to highlight key clusters of viable CD19^+^ lymphocytes from a representative convalescent donor with an enriched memory B cell subset. Heatmap overlay shows median expression for each target. **(b-f)** Healthy and convalescent donor cell frequencies for **(b)** IgM^+^ memory, **(c)** switched Memory, **(d)** CD11c^+^, **(e)** T-bet^+^, and **(f)** DN subsets among viable CD19^+^ lymphocytes. Bars represent mean ^+^/-SD. Multiple comparison analysis between each donor subgroup was done with Kruskal-Wallis test with Dunn’s correction. Adjusted p value was used to determine family-wise significance at alpha = 0.05. Healthy control *n* = 24, conv. total *n* = 40, asymp. *n* = 5, conv. early *n* = 12, conv. mid *n* = 6, conv. late *n* = 17, except for T-bet **(e)**, where *n* = 4, 9, 6, and 16, respectively.

Given the demonstrated role of CD11c^+^ T-bet^+^ B cells in responses to viral antigens (9, 10), and their presence during acute SARS-CoV-2 infection (4), we screened convalescent donor samples for the presence of CD11c^+^ T-bet^+^ B cells. The frequency of both CD11c^+^ and T-bet^+^ B cells were only slightly elevated in convalescent subjects compared to healthy controls, likely reflecting a return to B cell homeostasis **(Figure 3d-e)**. Likewise, significantly elevated frequencies of double-negative (DN) IgD-negative CD27-negative B cells were not observed in convalescent subjects **(Figure 3f)** in our cohort. These finding suggests that activated and DN populations may be preferentially involved in the early phase and/or critical clinical presentation of SARS-CoV-2 infection, as observed by Woodruff et al. (5) during acute severe COVID-19. These analyses reveal substantial heterogeneity not just in the B cell immunophenotype between subjects, but also within the B cell memory compartment itself.

### Shorter symptom duration was correlated with increased switched and IgM^+^ memory B cell frequencies in convalescent subjects

While we observed diverse B cell subsets in convalescent subjects, it was still unclear which, if any, of these subsets were correlated with clinical outcomes following symptomatic COVID-19 infection. An association between frequencies of memory B cells and enhanced recovery following COVID-19 pneumonia was reported (8). We therefore analyzed our B cell immunophenotyping dataset for its correlation with self-reported symptoms in our convalescent cohort. Our analysis revealed a significant negative correlation between the duration of COVID-19 symptoms and the frequency of memory B cells within the B cell compartment, as well as for the IgM^+^ memory B cell subset **(Figure 4a-b)**. A similar trend was observed for switched memory B cell frequency **(Figure 4c)**.

**Figure 4.**
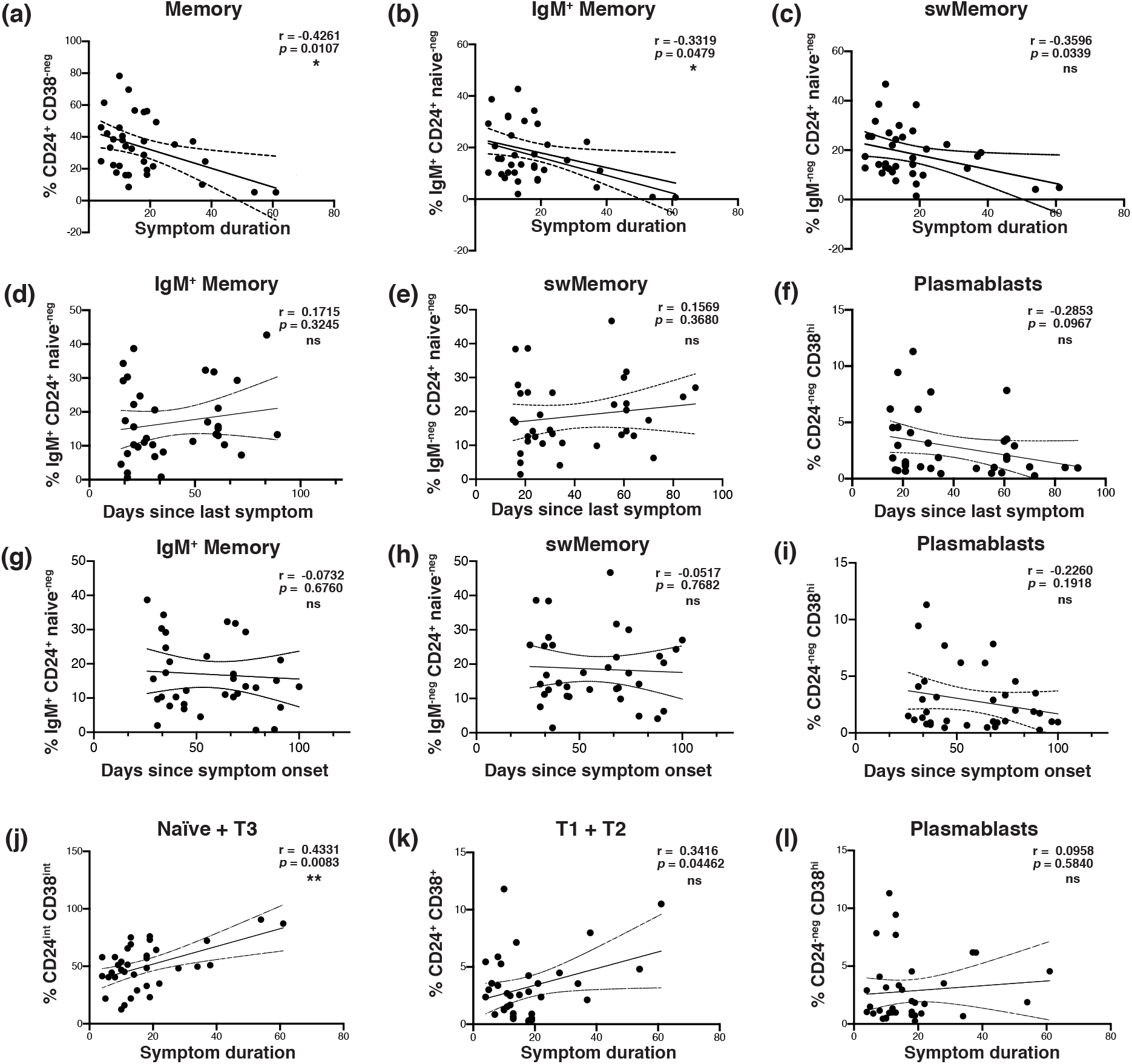
The frequency of memory B cells in the peripheral blood was correlated with shorter symptom duration following infection with SARS-CoV-2, and stable over time. **(a-c)** Scatterplot correlation of symptom duration in days, vs. the frequency among viable CD19^+^ B cells of **(a**) total memory, **(b)** IgM^+^ memory, and **(c)** switched memory. **(d-f)** Scatterplot correlation of days since last symptom vs. **(d)** IgM^+^ memory, **(e)** switched memory, and **(f)** plasmablast frequency among viable CD19^+^ lymphocytes. **(g-i)** Scatterplot correlation of days since symptom onset vs. **(g)** IgM^+^ memory, **(h)** switched memory, and **(i)** plasmablast frequency among viable CD19^+^ B cells. **(j-l)** Scatterplot correlation of symptom duration in days, vs. **(j)** naïve and transitional type 3, **(k)** transitional type 1 and 2, and **(l)** plasmablast frequency among viable CD19^+^ B cells. Pearson’s correlation r value and 95% confidence intervals shown with two-tailed *p* values, alpha = 0.05. *n* = 35 (all symptomatic subjects) for all plots.

To determine whether these observations were due solely to the time of sampling, we analyzed the frequency of B cell subsets in convalescent subjects and the number of days between last symptom and sample collection, as well as the number of days between symptom onset and sampling. IgM^+^ and switched memory B cell frequencies were stable or enhanced over time. We failed to observe a correlation between IgM^+^ or switched memory B cell frequency with the time since the last reported symptom **(Figure 4d-e)**, or the time since symptom onset **(Figure 4g-h)**. As was expected, longer symptom duration correlated with increased frequency of naïve and transitional B cells, due to contraction of the plasmablast response **(Figure 4g-k)**. Neither age nor gender were observed to have a statistically significant influence on any of the clinical or immunophenotypic parameters examined in this dataset (data not shown).

Several studies have reported robust expansion of peripheral blood plasmablasts during SARS-CoV-2 infection in some patients (5, 6). In agreement with these data, our cohort of convalescent subjects contained individuals with dramatically elevated frequencies of CD38^hi^ CD24-negative CD19^+^ plasmablasts **(Figure 1g)**. Despite this trend, this subset was not significantly correlated with the duration of COVID-19 symptoms at the time point at which we sampled **(Figure 4l)**. Plasmablast frequency among convalescent B cells did appear to wane over the time since last symptom, and symptom onset, confirming the contraction of the acute response **(Figure 4f, i)**. Collectively, these data suggest that the presence of B cell memory is a durable clinical correlate of shorter duration of COVID-19 disease.

### Memory B cell frequency was correlated with anti-RBD antibody production

We next addressed whether the frequency of B cell memory in convalescent subjects correlated with the generation of anti-spike receptor-binding domain (RBD) antibodies. The spike RBD is thought to be required for SARS-CoV-2 binding and entry via the ACE2 receptor, and both inhibitory and neutralizing anti-RBD antibodies have been identified in infected and recovered subjects (4, 11). In seropositive convalescent subjects, IgG1 and IgM anti-spike RBD were significantly correlated with CD24^+^ CD38-negative memory B cell frequency **(Figure 5a-b, f)**. This was not observed for IgG2, IgG3, or IgG4 **(Figure 5c-e**; for full cohort, see **supplemental Figure 5)**.

**Figure 5.**
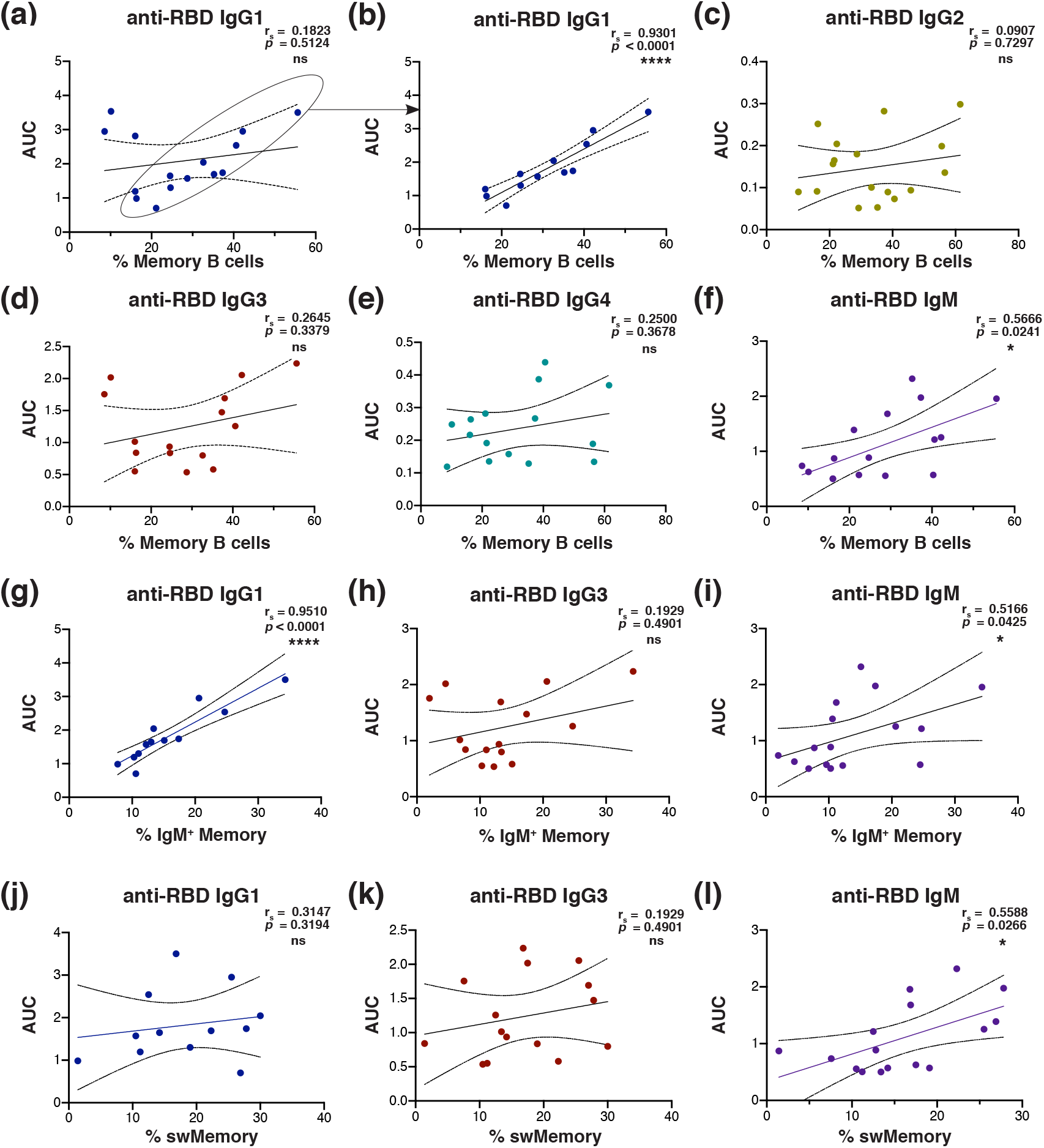
Anti-spike receptor binding domain antibody levels correlated with memory B cell frequency in convalescent plasma donors. **(a-f)** Scatterplot correlation of area under the curve for anti-RBD plasma absorbance vs. memory B cell frequency from convalescent subjects seropositive for each isotype (IgG1 & IgG3 *n* = 15, IgM *n* =16). Subjects whose data points are circled in **(a)** were isolated for statistical analysis of IgG1 in **(b, g, and j**; *n* = 12**). (g-i)** Scatterplot correlation of area under the curve for anti-RBD plasma absorbance vs. IgM^+^ memory B cell frequency among viable CD19^+^ lymphocytes. **(j-l)** Scatterplot correlation of area under the curve for anti-RBD plasma absorbance vs. switched memory B cell frequency among viable CD19^+^ lymphocytes. Spearman’s correlation r_s_ value and 95% confidence intervals shown with two-tailed p value, alpha = 0.05 for all analyses. Sample number distribution was the same for total, IgM^+^ and switched memory B cell analyses.

The positive relationship between IgG1 and memory held for IgM^+^ memory B cells **(Figure 5g)**, but not for switched memory **(Figure 5j)**. For almost all anti-RBD IgG1-producing subjects, there was a nearly dose-dependent correlation between and anti-RBD IgG1 and IgM^+^ memory B cell frequency. IgM anti-RBD was significantly correlated with both switched and unswitched memory, though not as markedly **(Figure 5i & l)**. Memory B cell frequency and total immunoglobulin levels were largely unrelated, except for a weak positive correlation between memory and total IgG2 **(supplemental Figure 5g-l)**.

### Memory B cell frequencies were maintained or increased as plasmablasts returned to baseline

We next addressed whether the memory B cells were a stable population, or waned, as did the plasmablast response. We re-sampled a subset of the convalescent cohort at least 3 months after the initial visit **(Figure 1, supplemental Figure 1)**. Results from this longitudinal analysis showed a contraction of the plasmablast response **(Figure 6a)**. We also observed that memory B cell frequencies and subset distribution were maintained or increased in most subjects **(Figure 6b-d)**. Naïve B cell frequency remained nearly constant **(Figure 6e)**, and transitional subset frequencies fluctuated over time, without exhibiting a trend **(Figure 6f)**. Although data from individual subjects demonstrated temporal variability with regards to the frequency of activation-associated B cell subsets, the frequency of T-bet^+^, CD11c^+^, DN, and activated B cells did not change significantly over time for the cohort as a whole **(Figure 6g-j)**. These trends were not obviously impacted by the stage of convalescence when the first sample was obtained.

**Figure 6.**
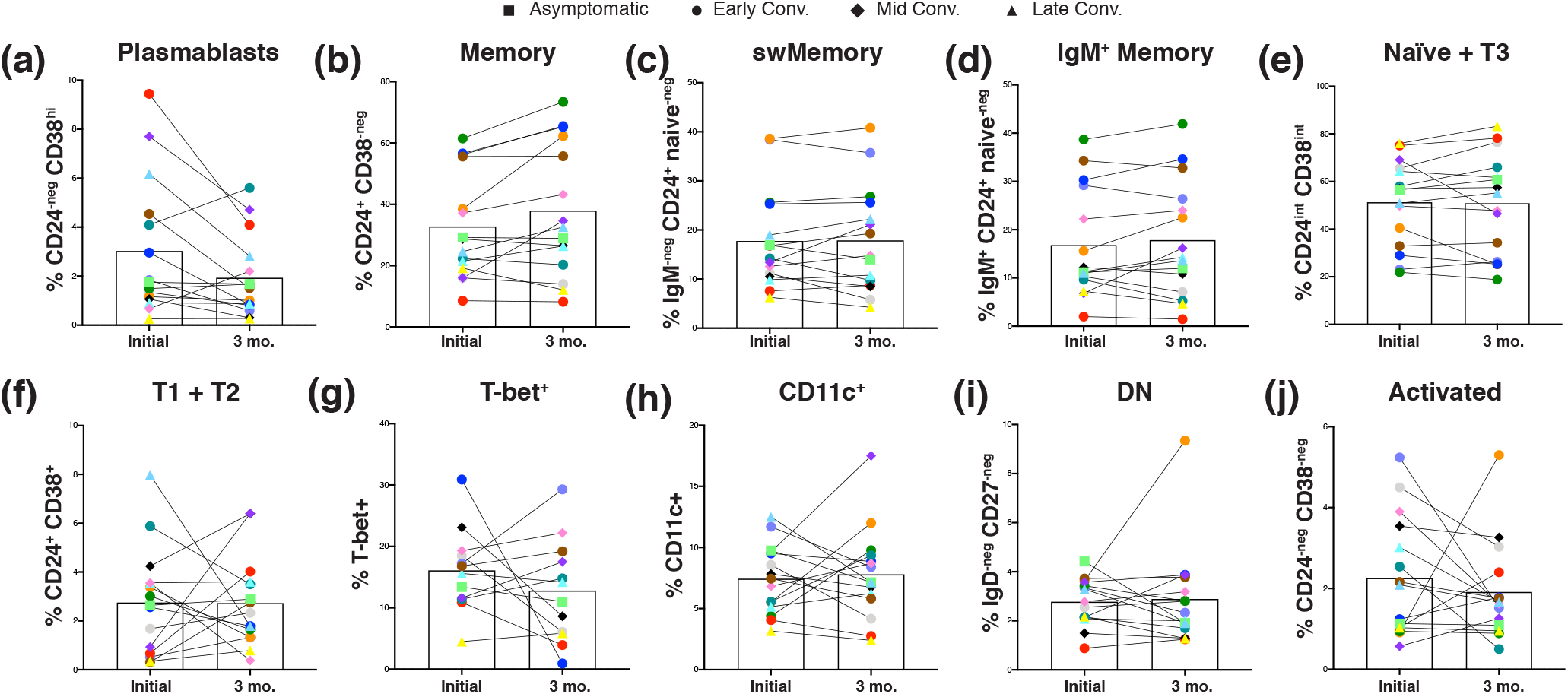
Convalescent SARS-CoV-2 subjects displayed a contraction of plasmablasts over time, but maintained memory B cells. **(a-j)** Frequency of **(a)** plasmablasts, **(b)** total memory, **(c)** switched memory, **(d)** IgM^+^ memory, **(e)** naïve ^+^ transitional 3, **(f)** transitional 1 + 2, **(g)** T-bet^+^, **(h)** CD11c^+^, **(i)** DN, and **(j)** activated cells among CD19^+^ viable lymphocytes from 15 convalescent plasma donors at initial draw and 3-month follow-up visit. Each pair of connected points (and color) represents an individual subject. Symbol shapes indicate convalescent subset. Bars represent mean. Two donors (one asymp., one late conv.) were omitted due to B cell abnormalities likely unrelated to SARS-CoV-2.

None of the B cell subsets analyzed exhibited statistically significant changes over the 3-month period wherein we assessed our convalescent cohort, although the plasmablast compartment contraction suggests a return to B cell homeostasis. These findings support a role for switched and unswitched memory B cells in the maintenance of stable, durable SARS-CoV-2 immunity.

## Discussion

In this study we identified both unswitched and switched B cell memory as a correlate of shorter COVID-19 symptom duration. Moreover, IgM^+^ memory correlated strongly with the anti-RBD IgG1 antibody response. These data suggest that a protective memory response occurs in at least some COVID-19 patients, preceded or accompanied by the generation of IgM^+^ memory B cells. We envision three possible explanations for these findings. First, it is possible that memory B cells identified in some individuals were generated in response to a previous coronavirus infection. Coronaviruses as a group likely generate cross-reactive B and T cells responses (12). The observation that the anti-RBD IgG1 response was correlated with IgM^+^ memory cell frequency is paradoxical, however, given that IgM^+^ memory cells don’t produce switched immunoglobulin. We propose that IgM^+^ memory cells are generated in abundance during coronavirus infections, and that some of these enter germinal centers and undergo class switching following a related coronavirus infection, thereby contributing to enhanced IgG1 production. The capacity of IgM^+^ memory cells to preferentially enter germinal centers upon activation has been well-documented in mouse and human studies (13, 14, 15). This characteristic versatility of IgM^+^ memory cells could be advantageous for immunity to pathogens, such as the coronaviruses, where infections with closely related strains often occur.

The lack of correlation between the frequency of resting memory B cells and CD11c^+^ and/or T-bet^+^ B cells in convalescent subjects was unexpected, given the pivotal role these molecules play in type-1 B cell immunity. We hypothesize that these factors may play a key role during the acute phase and during chronic viral infections, but are not essential during the convalescent phase of SARS-CoV-2 infection. Additional prospective studies and kinetic analyses of previously-exposed and naïve individuals will help to resolve this question.

A second explanation for the enhancement of COVID-19 recovery coincident with memory B cell expansion, is that naïve subjects whose B cells received more efficient T cell help during primary infection generated a larger pool of memory B cells. This explanation is consistent with the close relationship between memory and pathogen-specific antibody production we observed. This explanation would suggest that T cells contributed to a better germinal center response in some individuals. In contrast, subjects whose B cells received insufficient or inappropriate T cell help would generate poor germinal center reactions, fewer antigen-specific antibodies, and have a longer symptomatic disease period.

T cell help occurs largely within germinal centers, suggesting that the local immune environment may be critical for an adequate response to SARS-CoV-2. Indeed, severe disruption of lymphatic tissue organization and germinal center formation have been observed in severe COVID-19 cases (16). Moreover, studies from our laboratory have shown that follicle architecture disruption, acute plasmablast expansion, and type-1 cytokine skewing occurs during murine intracellular bacterial infection (17-19). We observed IgM^+^ memory to be protective during *E. muris* infection, despite severe immunopathology associated with primary infection. In this way, it is possible that the germinal center disruption caused by SARS-CoV-2 infection also contributes to the generation of protective IgM^+^ B cell memory. The data herein suggests that IgM^+^ memory B cells may complement long-lived plasma cells generated via germinal centers and provide protection against subsequent SARS-CoV infection.

Finally, the benefit of memory B cell expansion may only be an indication that acute inflammation and excessive cytokine production did not occur in some individuals. Under inflammatory conditions, such as those occurring in systemic lupus erythematous (SLE), B cell subsets have been shown to be skewed toward an activated, extrafollicular effector fate (20). It has been suggested that B cell responses to severe acute COVID-19 disease have similar characteristics to those observed in SLE (5). The variability in the penetrance of this phenotype may be explained by the influence of gender and pre-existing autoimmunity. Future work using animal models of COVID-19 may help to resolve these questions.

Our studies support other work that has reported that germinal center-derived memory B cells are likely generated following SARS-CoV-2 infection, and that these memory B cells are a durable correlate of an effective primary response (4, 8). These studies challenge early claims of waning immunity shortly after SARS-CoV-2 infection. Our data also show that some individuals can be identified as having better natural immunity. We also propose that both IgM^+^ and switched memory B cells may provide a good indication of vaccine efficacy, and that individuals with large numbers of IgM^+^ memory B cells may be better protected from future re-infection with homotypic or heterotypic infection. Our findings suggest that IgM^+^ memory B cells are central to the COVID-19 adaptive immune response, and highlight the need for prospective SARS-CoV-2 studies to determine whether large memory B cell populations are a pre-existing correlate of protection, or a durable measure of the antiviral immune response.

## Methods

### Study Design

Study participants were recruited at the SUNY Upstate Medical University Clinical Research Unit starting from March 2020 and is ongoing. Participants meeting eligibility criteria were adults aged 18 or older who have tested positive for SARS-CoV-2 and are at least 14 days past their first symptom. Exclusion criteria included the inability to give informed consent and/or an inability to donate plasma or blood transfusion in the past. Study participants were interviewed by study staff, and presented to the SUNY Upstate Clinical Research Unit for peripheral blood collection. Information regarding symptoms, including dates of first and last symptom, was self-reported. Donors were questioned about acute symptoms such as fever, shortness of breath, sore throat, cough that impacted activity, and fatigue that impacted activity. These indications were used to calculate dates of symptoms retrospectively. Lingering symptoms such as loss of taste and smell, mild cough or tickle in the throat, or lingering fatigue that did not impact their daily activity were not considered part of the acute illness and therefore not included in the length of illness. For donors reporting no symptoms, the date of positive RT-PCR test was used for the start and stop date of symptoms. These subjects were not included in correlative analysis of symptom duration. Healthy control subjects were adults aged 18 or older who denied infection with or known exposure to SARS-CoV-2. Healthy controls were screened by anti-RBD plasma ELISA to confirm negative exposure status. Sample size was determined based on subject availability. All samples were de-identified following collection, and researchers conducting assays were blinded to clinical data until final comparative analysis.

### Blood sample processing and storage

PBMCs were obtained following gradient centrifugal separation of peripheral blood using Cell Preparation Tubes (CPT) (BD) for 30 minutes at 1700 x g. Plasma was separated, aliquoted, and stored at -20°C for antibody assays. The mononuclear layer was washed in PBS prior to counting on a Coulter particle counter (Becton-Dickinson). PBMCs were either directly stained for flow cytometry (initial visit samples), or frozen slowly to -80°C in FBS and DMSO for short-term storage (3-month visit samples). Flow cytometry panel was validated using a sample of fresh vs. frozen PBMCs to ensure comparability in target detection. Plasma samples were heat-inactivated (56°C for 30 minutes) prior to use in assays.

### Flow Cytometry

The following antibodies used for flow cytometry were obtained from BioLegend: CD21 (Bu32), T-bet (4B10), CD38 (HIT2), CD11c (Bu15), CD3 (HIT3A), CD14 (HCD14), IgD (IA6-2), CD24 (MC5), IgM (MHM-88), CD27 (O323), and CD19 (HIB19). For flow cytometric analysis, single-cell suspensions were incubated with 1 microg/ml anti-CD16/CD32 in 2% normal goat serum/HBSS/0.1% sodium azide (Fc blocking solution). The cells were then stained with aqua Live/Dead stain (Invitrogen) and washed prior to incubation with mAbs. Cells were fixed, permeabilized, and stained for intracellular targets using an intracellular staining kit (BD). Unstained controls were used to set the flow cytometer photomultiplier tube voltages, and single-color positive controls were used to adjust instrument compensation settings. Data from stained samples were acquired using a BD Fortessa flow cytometer equipped with DIVA software (BD Biosciences) and were analyzed using FlowJo software (Tree Star). t-SNE visualization was generated using FlowJo automatic (opt-SNE) learning configuration, with 1000 iterations, a perplexity of 30, learning rate of 3179, exact KNN algorithm, and Barnes-Hut gradient algorithm.

### ELISA protocol

Plasma samples were first heat-inactivated at 56°C for 30 mins before use in assays. Recombinant Twin-Strep-tagged RBD protein was purified from 293T cells transfected with paH-RBD SD1-3CH25, generously provided by Jason S. McLellan (University of Texas), as previously described (21). RBD protein was coated on Strep-Tactin® microplates plates (IBA, Germany) in binding buffer (100mM Tris pH 8, 150mM NaCl, 1mM EDTA) overnight at 4°C. Plates were washed three times with PBS-T (1x PBS/0.05% Tween-20) and subsequently blocked with 3% BSA (Sigma) in PBS-T for 1 hour at RT. Diluted plasma was loaded onto plates and incubated for 2 hours at RT. After incubation, plates were washed three times with PBS-T and HRP-conjugated secondary anti-human antibodies were used for detection. Total IgG was detected using goat-anti-human-IgG-HRP (Rockland, #209-1304). IgM was detected using goat-anti-human-IgM-HRP (Sigma, #A6907), and IgG subclass antibodies were detected with mouse horseradish peroxidase (HRP)-conjugated anti-human IgG1 (9054-05), IgG2 (9060-05), IgG3 (9210-05), and IgG4 (9200-05) from SouthernBiotech. Wells were washed three times with PBS-T before the addition of HRP substrate SIGMAFAST OPD (Sigma, #P9187). Reaction was quenched with 1M hydrochloric acid (Fisher Scientific, #S25856) and read at 490 nM on a BioTek Synergy LX multi-mode plate reader. Area under the curve (AUC) analysis was performed using peak identification set to a 10% minimum change from baseline. Plasma ELISAs for total human immunoglobulin isotype quantitation were performed using the Human Immunoglobulin Isotyping LEGENDplex 6-plex kit (BioLegend) according to the manufacturer’s instructions. Data were collected using a BD LSR II flow cytometer and analyzed using LEGENDplex Data Analysis Software.

### Statistical Analysis

Statistical analyses were performed using GraphPad Prism software (v8.4.3). Analysis of correlation between flow cytometry, total serum immunoglobulin ELISA data, and continuous clinical data was performed using Pearson correlation coefficients for data sets equal to or larger than 35 values, or nonparametric Spearman’s Rank correlation for data sets with fewer than 35 values. p-values are two-tailed and 95% confidence intervals shown where noted in figure legends. Statistical analysis of cell subset frequency between healthy and the total convalescent donor cohort from flow cytometry assays was performed using unpaired nonparametric Mann-Whitney test with two-tailed p-values and 95% confidence intervals. Multiple comparison analysis between each convalescent subject subgroup was done with Kruskal-Wallis test with Dunn’s correction. Adjusted *p* value was used to determine family-wise significance at alpha = 0.05. Statistical analysis of anti-RBD plasma ELISA data was performed using area under the curve analysis with peak identification set to a 10% minimum change from baseline. NS indicates a *p* value > 0.05, *p, < 0.05, **p, < 0.01, ***p, < 0.001, and ****p, <0.0001. The statistical tests performed are indicated in the figure legends. For column graphs, the column in each plot indicates the arithmetic mean of the dataset, and upper and lower bounds indicate SD of the dataset.

### Study approval

All participants provided written informed consent prior to participation in the study, which was performed according to a protocol approved by the Institutional Review Board (IRB) of the SUNY Upstate Medical University under IRB number 1587400. All clinical investigation was conducted according to Declaration of Helsinki principles.

### Author contributions

K.L.N., H.E.T., J.R.W., and G.M.W. conceived, designed, and analyzed the experiments. T.P.E. designed clinical protocols. K.L.N. and J.B.C. performed flow cytometry. D.C.C., Y.I.K. and H. E.T. performed anti-RBD ELISAs, and V.Z.R. carried out total plasma Ig ELISAs. T.P.E. recruited participants and executed clinical protocols. K.L.N. and J.B.C. processed clinical samples. K.L.N. authored the manuscript, with editing by J.R.W., H.E.T., T.P.E., and G.M.W. All authors reviewed and approved the manuscript.

## Data Availability

All data generated or analyzed during this study are included in this article and supplementary data, or available from the corresponding author upon reasonable request.

## Acknowledgments

We thank all study participants who made this work possible, the SUNY Upstate Medical University Clinical Research Unit, Michelle Klick for research coordination, the SUNY Upstate Institute for Global Health and Translational Science for research administration, Dongliang Wang for assistance with statistical analysis, Mark Abbott and Kristen Baxter for critical laboratory support, Lisa Phelps and Steven Taffet for support with the SUNY Upstate Medical University flow cytometry core, and Russell Levack for helpful discussion. This work was supported by U.S. Department of Health and Human Services Grant R01AI114545 (G.M.W.), SUNY Office of Research and Economic Development seed grant RFP #20-03-COVID 202077 (J.R.W. and G.M.W.), and internal funding from SUNY Upstate Medical University.

